# Automated recognition of ultrasound cardiac views based on deep learning with graph constraint

**DOI:** 10.1101/2020.05.07.20094045

**Authors:** Yanhua Gao, Yuan Zhu, Bo Liu, Yue Hu, Youmin Guo

## Abstract

**Objective:** In Transthoracic echocardiographic (TTE) examination, it is essential to identify the cardiac views accurately. Computer-aided recognition is expected to improve the accuracy of the TTE examination.

**Methods:** This paper proposes a new method for automatic recognition of cardiac views based on deep learning, including three strategies. First, A spatial transform network is performed to learn cardiac shape changes during the cardiac cycle, which reduces intra-class variability. Second, a channel attention mechanism is introduced to adaptively recalibrates channel-wise feature responses. Finally, unlike conventional deep learning methods, which learned each input images individually, the structured signals are applied by a graph of similarities among images. These signals are transformed into the graph-based image embedding, which act as unsupervised regularization constraints to improve the generalization accuracy.

**Results:** The proposed method was trained and tested in 171792 cardiac images from 584 subjects. Compared with the known result of the state of the art, the overall accuracy of the proposed method on cardiac image classification is 99.10% vs. 91.7%, and the mean AUC is 99.36%. Moreover, the overall accuracy is 98.15%, and the mean AUC is 98.96% on an independent test set with 34211 images from 100 subjects.

**Conclusion:** The method of this paper achieved the results of the state of the art, which is expected to be an automated recognition tool for cardiac views recognition. The work confirms the potential of deep learning on ultrasound medicine.

## Introduce

Transthoracic echocardiogram (TTE) is the most commonly used cardiac examination tool, which provides comprehensive observations on the cardiac structures and functions, and assist in the diagnosis and guide the treatment [1] [2], TTE have a wide range of applications [3] [4], The American Society of Echocardiography (ASE) and has established the standards for two-dimensional(2D) TTE examinations [3] [5]. Cardiologists and echocardiographers must follow these standards, which not only ensures that pathological changes are not missed, but also helps ensure that the diagnosis is repeatable.

These 2D ultrasound cardiac views provide qualitative imaging of the heart from multiple angles, as well as enough information about the structures of the heart, such as the ventricles, leaflets and annulus, pericardium and great vessels. However, the accurate scan of the cardiac view is closely related to the operators’ experiences [6], Lots of reasons lead to the increase of diagnostic variability [7]. Due to the non-standard views, even if the left ventricle is carefully tracked, the shortened left ventricle will lead to an underestimation of the maximum volume. TTE usually requires echocardiographers with a lot of experiences, besides, it is a complex and time-consuming. Meanwhile, all the correct diagnosis depend on accurate identification of cardiac views.

The main challenge of ultrasound medicine is the low image quality, which is caused by noise and artifacts. The computer-aided analysis of ultrasound images has received widespread attention. Deep learning has been applied to ultrasound image analysis in recent years [8] [9], Ultrasound image classification provides effective decision support tools including the identification of tumors and lesions, such as breast cancer and benign lesions[10] [11] [12], liver cancer[13] [14] and Thyroid nodules[15] [16], The other applications are the fetuses and neonates, including quality control of fetal ultrasound and standard views of the fetus [17] [18] [19] [20],

Detection of regions of interest (ROI) is another interesting task. Segmentation of tissue structure and lesions helps quantitative analysis of clinical parameters, including non-rigid organ segmentation [21] [22] and rigid organ segmentation [23] [24], 3D ultrasound image analysis has not been widely used because of expensive calculations and limited data volume [25] [26],

The accurate identification of cardiac cycle (cardiac cycle phases, end-diastolic and end-systolic) is a prerequisite for the measurement of cardiac physiological parameters, such as stroke volume, ejection fraction, and end diastolic volume. Dezaki provided a deep residual recurrent neural network to automatically identify cardiac cycle phases [27]. Sofka provided a fully convolutional regression network to detect the measurement points of the parasternal long axis view [28], The combination of reinforcement learning and deep learning is used for anatomical (cardiac US) landmark detection [29], Madani proposed a fast and accurate cardiac view recognition method based on convolutional neural networks, which achieved the known excellent accuracy of 91.7% (image classification) and 97.8% (video classification) [30].

The challenge of automatic recognition of cardiac views comes from large intra-class differences and small inter-class differences. First, the individual differences of people such as gender, race, age and so on. Second, heart disease leads to the changes of cardiac shapes, for example the hypertension causes left ventricular enlargement and wall thickening. Third, during the cardiac cycle, the cardiac surface changes periodically and non-linearly. Fourth, the scanning parameters, such as different magnifications and signal gains, may cause great changes of ultrasound images. These factors cause large intra-category differences, and even experienced echocardiographers may not be able to identify standard cardiac views accurately enough. see Supplementary Figure 1.

This paper proposes an automatic recognition method to identify nine conventional cardiac views, i.e. Parasternal Long-axis (PSLA), Parasternal short-axis at the level great vessels(basal short axis, sax-basal), Parasternal short-axis at the level of papillary muscles or mitral (short axis at mid, sax-mid), Apical Four-Chamber view (a4c), Apical Five-Chamber view (a5c), Apical Two-Chamber view (a2c), Apical Three-Chamber view (a3c), Subcostal Four-Chamber view (sub4c), Suprasternal notch Aortic arch (supao). These views are included in a standard ultrasound cardiac examination. The presented method is based on convolutional neural networks(CNN), which includes three effective strategies, i.e. graph regularization learning [31], combined with spatial transform networks [32] and channel attention mechanism [33], The highlights are given as follows.

1. Unlike conventional deep learning methods, which learned each input images individually, the structured signals are applied in neural network training by a graph of similarities between images. The structured signals represent the relationships or similarities between input images. This signal is transformed into graph-based image embedding, which acts as unsupervised regularization constraint to improve the generalization accuracy [31].
2. Considering the cardiac views have large shape changes during cardiac cycle, the spatial transformer network is performed as an independent pre-processing module, which learns the deformation during cardiac cycle. Second, the channel attention mechanism (squeeze-and-excitation network) is introduced to adaptively recalibrates channel-wise feature responses to enhance channels that have a significant impact on the identification.

Our method achieved state-of-the-art performance on two independent datasets. The presented method is expected to be an assistant diagnostic tool for daily cardiac examinations.

## Method

### Datasets

All the cardiac images came from two hospitals, Shaanxi Provincial People’s Hospital (SXPPH) and the First Affiliated Hospital of Xi’an Jiaotong University(XJTUFAH). Four experienced echocardiographers recorded the videos. They are no longer involved in the next phase of research. we recruited 584 subjects and 100 subjects from SXPPH and XJTUFAH respectively, and each echocardiographic subject included the video images of 9 cardiac views. More information refers to the supplementary Table 1 and 2.

### Preprocessing pipeline

In order to remove the patients’ private information, the surrounding pixels of each video were cut out. An image was extracted from the videos at an interval of 5 frames. Approximately 200-400 images were obtained from each video. All images were reviewed independently by two experienced cardiac ultrasound doctors, the low-quality and incorrect images were excluded, and only the images agreed by both doctors were retained. Finally, 171792 (SXPPH, Dataset1) and 34211 (XJTUFAH, Dataset2) images were obtained. The data distribution is shown in Table 1 and Table 2. clinical information is shown in supplementary Tables 1 and 2. The study were approved by the consent of the Institutional Review Board (IRB), and the subjects were informed of experiment content and risks.

**Table 1.** the distribution of cardiac views from SXPPH

| Cardiac views | PSLA | SB | SM | a4c | a5c | a2c | a3c | sub4c | supao | Total amount |
| --- | --- | --- | --- | --- | --- | --- | --- | --- | --- | --- |
| amount | 27888 | 25355 | 28546 | 15433 | 18270 | 14760 | 17099 | 12988 | 11455 | 171792 |

**Table 2.** the distribution of cardiac views from XJTUFAH

| Cardiac views | PSLA | SB | SM | a4c | a5c | a2c | a3c | sub4c | supao | Total amount |
| --- | --- | --- | --- | --- | --- | --- | --- | --- | --- | --- |
| amount | 4162 | 5140 | 4205 | 3113 | 3783 | 5086 | 3283 | 2886 | 2553 | 34211 |

The images in dataset 1 were divided into a training, validation, and test set according to the ratio of 7:1:2. The images from the same subject were not divided into different sets for the data independence. The dataset2 was used as an independent test set to confirm generalization accuracy between different hospitals. All images were scaled to the 512×512 pixels and RGB channels to meet the following network framework.

### Network framework

A graph-constrained convolutional neural network (CNN) framework was proposed. The graph is built based on the similarity among images. Each node on the graph represents a training image, and the edge weights between every two nodes indicates the similarity between two images represented by the two nodes. The learning strategy is based on the assumption that more similar images are more likely to be the same labels. When an input image is fed into a neural network, the images of its adjacent nodes are also fed in the same batch. The image embedding of adjacent nodes could be used as a graph reguluarizer or unsupervised graph loss. Meanwhile, the cross-entropy of the label and the predicted probability of input image are then calculated as the supervised loss, as shown in Figure 1. The overall training goal is to minimize the weighted sum of supervised loss and unsupervised graph loss.

**Fig 1.**
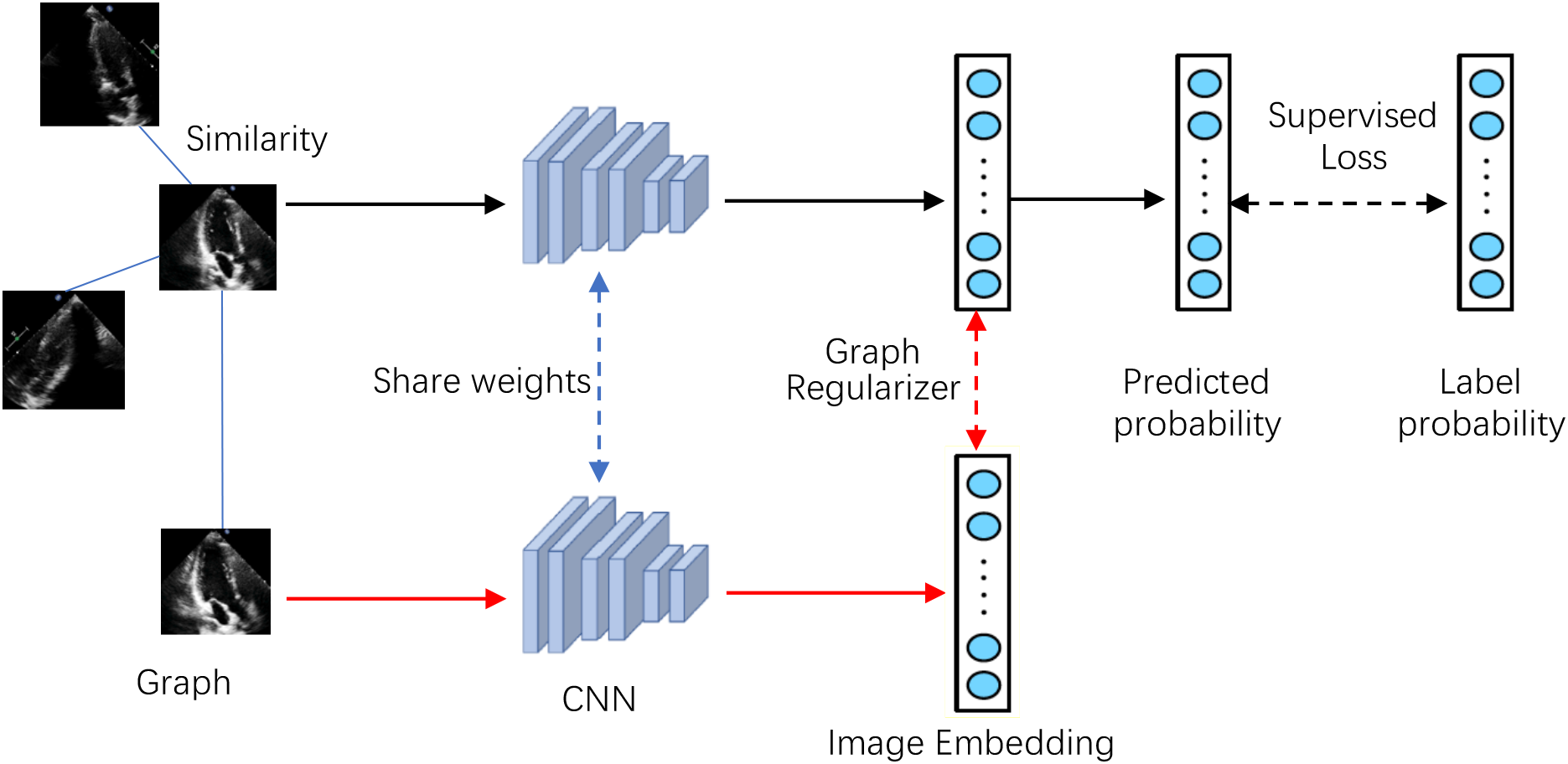
An illustration of the proposed classification framework.

The black flow represents the conventional CNN training. The red flow indicates that the adjacent images are fed in the same batch, and their image embedding is also computed for the graph regularizer, i.e. the graph loss. The cross-entropy of the true labels and predicted probabilities is the supervisor loss. The input image and its adjacent images share the weights of the CNN.

Total loss is given as follows:

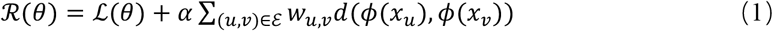

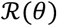 represents total loss, *θ* represents the weights of CNN. The first term 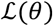 is supervised loss. The second term is the graph regularizer. *x_u_* is an input image. *w_u,v_* represent the similarity between *x_u_* and its adjacent node *x_v_*, which is also the edge weight between them, *ϕ* represents the image representation extracted from the embedding layer, *d* is the distance metric function of the mean square errors of the two image embedding.

Our CNN network is consisted of three functional modules, namely the spatial transformer network (STM) [34], Inception-V3 [32] and squeeze-and-excitation Network (SE) [33]. Inception V3 was a famous CNN and has achieved excellent performance in many image classifications. Although Inception V3 has shown some translation invariance, it cannot handle the deformations during cardiac cycle phase well. The spatial transformer network uses a localisation network to learn the parameters of the geometric transformation and the transformed image is then fed to the Inception-V3. As a feature extractor, the feature maps with 2048 channels from inception-V3 are fed into the SE. SE introduces the channel attention mechanism, which enhances the channel of feature maps that are more effective for accurate predictions. The detailed information is shown in supplementary Figure 3.

### Graph construction

During cardiac cycle, the ultrasound images are always changing, but they are similar. We used mutual information to represent their similarity. It is defined as follows.

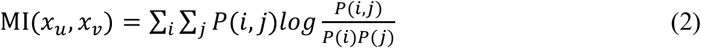

Because the category in the training sets is known, the mutual information is calculated for arbitrary two images of the same category. When It is greater than a threshold, two images are considered similar, and the mutual information is used as their edge weights. In order to control the graph size, the threshold was set to 0.2, and each node had a maximum of 10 adjacent nodes.

### Training process

The weights of STM and SE network were initialized with the glorot_uniform initializer. The Inception V3 was initialized by the pre-trained weights on ImageNet and deep fine-tuning was then performed. Because the image and the images of the adjacent nodes are loaded together in a batch, the number of images is limited by the GPU memory. For 32GB GPU, the batch size can be set to be 32, where the number of input images is 8, and the 4 adjacent images of each image are randomly selected. Base on the hyperparameter selection, the optimizer was set to Adam, the training epoch was 500, with 100 steps in each epoch. The learning rate was initialized to 0.001, with exponential function decrease by ratio 0.9 in every 5000 steps. When the validation accuracy did not improve for 5 epochs, the training was stopped. We compared the performance of *α* in formula (1), and found *α* = 0.4 can get the best results. During training, we did not use data augmentation, because the calculation of the graph would be greatly increased.

## Results

The two datasets from two hospitals include 171792 images (Table 1) and 34211 (Table 2). The cardiac views are involved in PSLA, sax-basal (SB), sax-mid (SM), a4c, a5c, a2c, a3c, sub4c, supao. Firstly, the training and testing are performed on the dataset1, and then independent test are performed on the dataset2.

Our proposed network is consisted of three functional modules, namely the spatial transformer network (STM), Inception-V3 and squeeze-and-excitation Network (SE). In order to verify the performance of the proposed framework, the baseline network was set to Inception V3. Inception V3 + SE, STM + Inception V3 + SE network, and Graph + STM + Inception V3 + SE network are then tested. In Inception V3, Inception V3 + SE, STM + Inception V3 + SE network training, the batch size was set to 32, and data augmentation is used, such as random flips, rotations, etc. We select the optimized hyperparameters based on validation sets.

The results are evaluated with the metrics such as the accuracy, sensitivity, specificity, and AUC. All metrics are calculated separately in a single category (cardiac view), defining the current category as positive class, and the other 8 categories are defined as negative classes. The accuracy is defined as the number of correctly classified samples divided by the number of all samples, and the sensitivity is defined as the number of correctly classified positive samples divided by the number of all positive samples, specificity is defined as the correctly classified negative samples divided by all the negative samples. The AUC is the Area Under Curve, which describes overall performance of sensitivity and specificity. Finally, the overall accuracy was calculated over all 9 categories. The overall accuracy of four networks and reference [30] is given as follows:

**Table 3.** Accuracy of four models in dataset1

| Networks | Overall accuracy |
| --- | --- |
| Inception V3 | 88.78% |
| Inception V3+SE | 94.65% |
| STM+Inception V3+SE | 96.50% |
| Graph+ STM+Inception V3+SE | 99.10% |
| Reference[30] | 91.7% |

The accuracy of Inception V3 network was only 88.78%. After channel attention introduced, the overall accuracy of Inception V3 + SE is improved significantly. STM reduces the variability of cardiac deformation, and slightly improves the accuracy to 96.50%. Because the graph regularization serves as a robust unsupervised loss, the proposed method achieves the best overall accuracy 99.10%, better than reference [30], the known excellent result.

The evaluation on cardiac views is shown in Table 4. PSLA, sax-basal, sax-mid, sub4c and supao are all recognized, with the sensitivity of 100%, and no images are misclassified into other categories, and the AUC reaches 100%. The a4c, a5c, a2c, and a3c are slightly misclassified. In particular, the sensitivity of a2c is only 94.63%.

**Table 4.** Test results of the presented method on dataset1. a: the mean of AUC

| Cardiac view | Sensitivity | specificity | accuracy | AUC |
| --- | --- | --- | --- | --- |
| PSLA | 100% | 100% | 100% | 100% |
| Sax-basal | 100% | 100% | 100% | 100% |
| Sax-mid | 100% | 100% | 100% | 100% |
| a4c | 100% | 99.82% | 99.84% | 99.91% |
| a5c | 97.78% | 100% | 99.74% | 98.89% |
| a2c | 94.63% | 99.47% | 98.94% | 97.05% |
| a3c | 97.78% | 99.16% | 99.00% | 98.47% |
| sub4c | 100% | 100% | 100% | 100% |
| supao | 100% | 100% | 100% | 100% |
| Total views | - | - | 99.10% | 99.36%<sup>a</sup> |

The evaluation on independent test set are shown in table 5. The Parasternal Long Axis, sax-basal, sax-mid, sub4c, and supao were all correctly identified. However, few images of the supao and sax-mid are mistakenly classified. Similarly, some images of a4c, a5c, a2c, a3c are misclassified. In particular, the sensitivity of a2c is reduced to 90.73%.

**Table 5.**
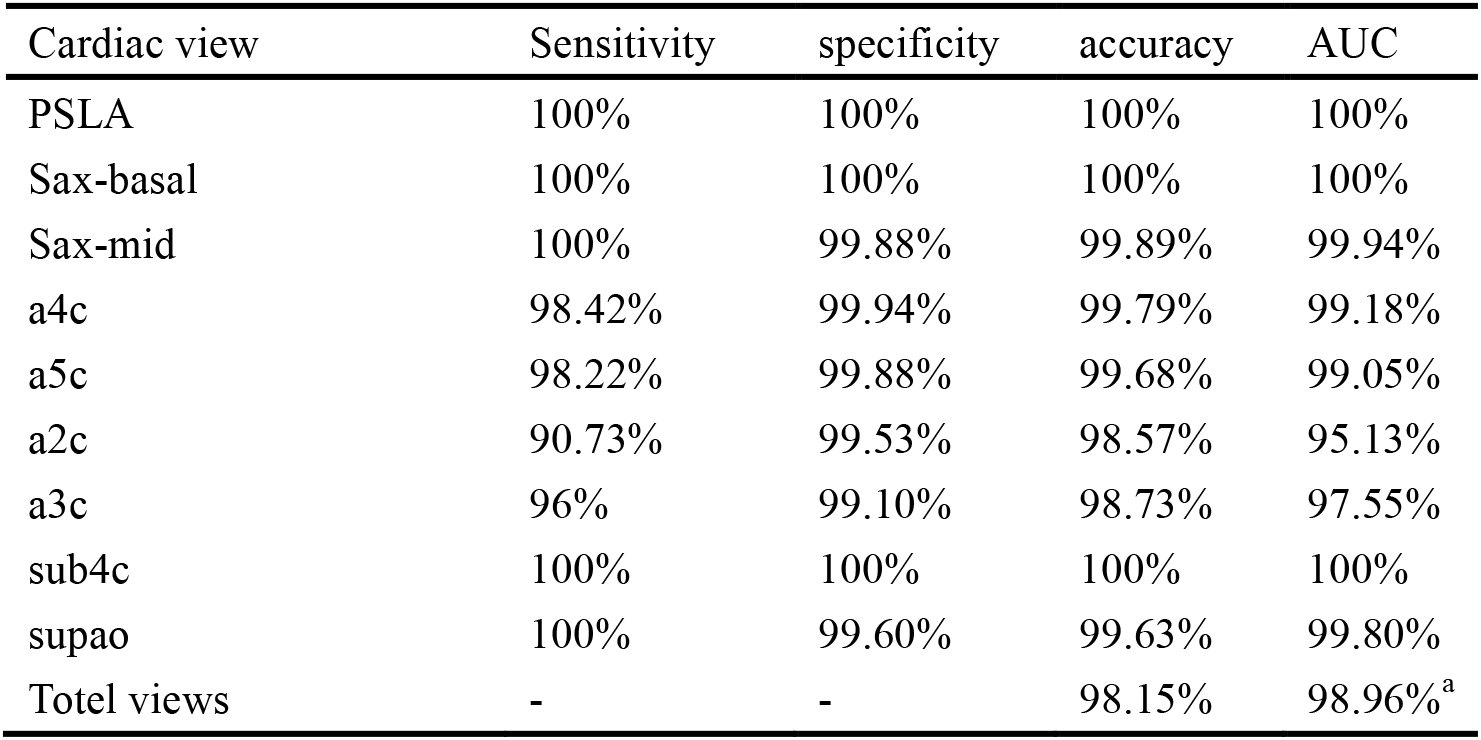
Test results of the presented method on dataset2. a: the mean of AUC

The overall accuracy of each category is all higher than 98%, and the AUC is more than 97%. The results in table 5 was only slightly worse than those of Table 4, but good generalization on different data sets are shown.

In order to find the classification errors among cardiac views, confusional matrices are computed. As shown in figure 2, the horizontal axis is the true labels, and the vertical axis is the predicted labels. The numbers on the Figure 2 are the percentages. On the diagonal, the closer the number is to 100, the more accurate the true and predicted labels are.

**Fig. 2.**
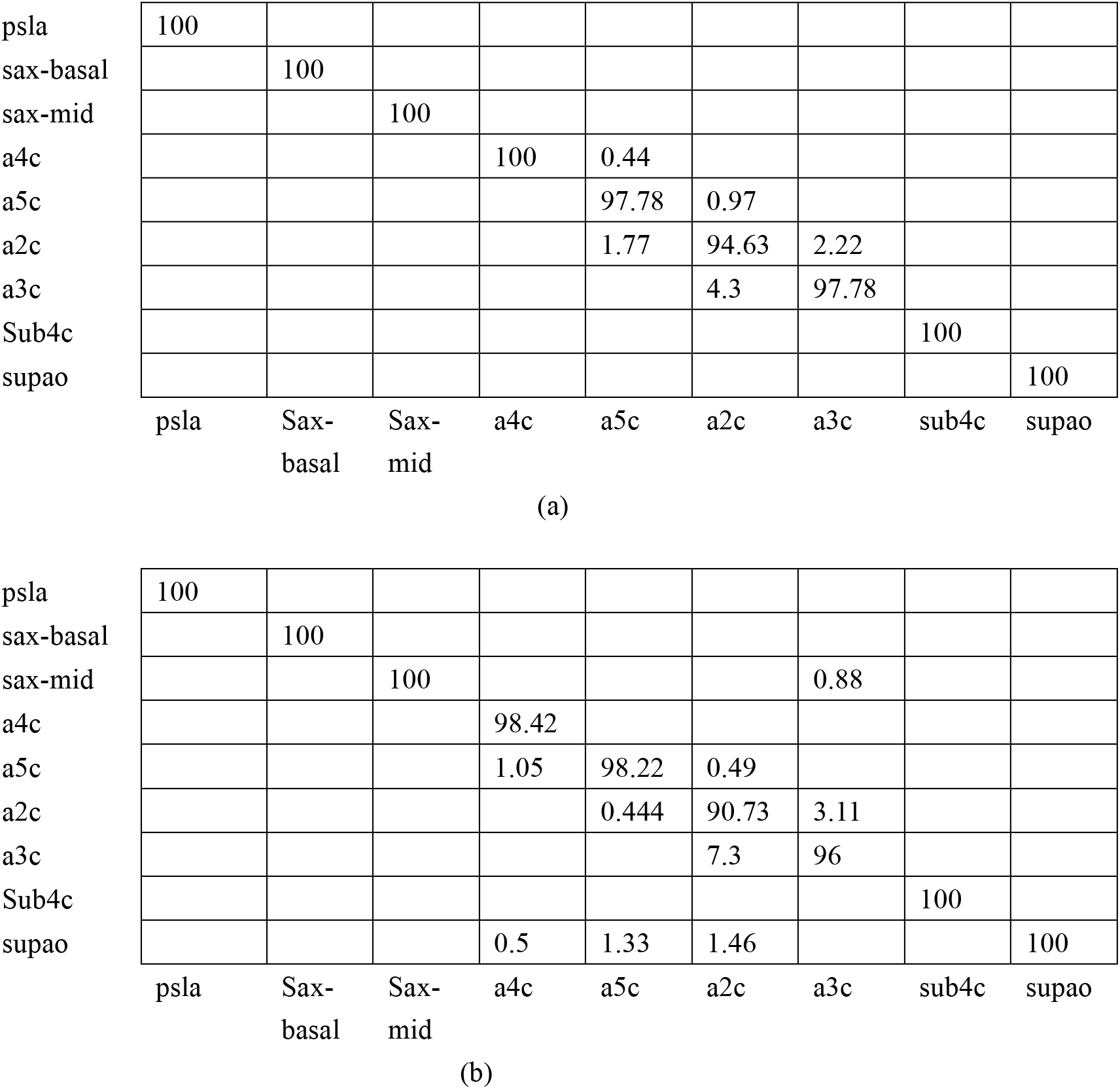
confusional matrices.

Figure 2(a) shows the confusion matrices of the test set in dataset1; Figure 2(b) is the confusion matrices in dataset2.

Each column is the ratio(percentage) of the predicted number of all categories divided by the total number of the category expressed by the column. The diagonal is the proportion of correct predictions for each category, and 100 means all samples are predicted correctly.

The confusion matrices show the misclassification among categories. The classification of PSLA, sax-basal, sub4c, supao, and a4c were accurate enough. However, the misclassification mainly occurs among a5c, a2c, and a3c. In particular, a2c and a3c are easily confused. In Fig2 (b), about 7% of a2c is misclassified as a3c, and a3c is 3.11 misclassified as a2c.

The visualization analysis based on t-SNE cluster is given in supplementary Figure3. After deep learning, the PSLA, sax-basal, sax-mid, sub4c, and supao were completely distinguishable. Only a few samples of a4c, a5c, a2c, and a3c are mixed together. The t-SNE clusters and the results of confusion matrix are consistent. An occlusion experiment is also shown in supplementary Figure 4, and the results indicate that our method can find important heart tissues in images.

## Discussion

The Echocardiography is still one of the most important cardiac examination because it is non-invasive, cost-effective, convenient. The diagnosis of heart diseases relies on the accurate identification of cardiac views. However, it depends on operators’ experiences, and the training is a complex and time-consuming process. Artificial intelligence is a good solution, which improves identification accuracy and reduces burdens of sonographer.

The main challenge of recognition for cardiac views is low-quality images, and great image change during the cardiac cycle. We used Inception V3 as the baseline network, but its overall accuracy was only 88.78%. SE network was then introduced which recalibrates channel-wise feature responses that were more effective for recognition through channel attentions. STM was also used to reduce the shape differences during cardiac cycle. It was worth noting that STM models the geometric deformation by 2D affine transform, which was a simplified version of the actual deformations. However, STM reduced the impact of the cardiac cycle on the recognition effectively. The accuracy was increased to 96.5%. To our knowledge, this result was better than the best known result [30].

Unlike conventional deep learning, the structural signals are introduced by the similarity between samples to learn relationships among them. Ideally, graph regularization can reduce the amount of labelled data and generalization errors. The first step of graph regularization is to build a graph. In general, the similarity between two images is not easy to be evaluated based on pixel-level comparisons. The cardiac images in the same cardiac cycle are similar and appear periodically. Therefore, the mutual information between two images can be used as a measurement of the edge weights. We introduced graph regularization to STM-inception V3-SE network, which further improved the performance by about 2%.

The datasets came from nearly 700 patients of two hospitals. The echocardiographers had excellent skills on TTE, and they recorded all videos of cardiac views. In order to ensure the independence of subsequent study, two other sonographers reviewed all the images, excluded some unqualified images. We can believe that the data acquisition is reasonable.

Nine usual cardiac views were studied for automatic recognition. The overall accuracy of the four networks was tested, and it was confirmed that the presented method achieved the best accuracy, 99.15%. The sensitivity, specificity, accuracy, and AUC values were also calculated for the nine categories respectively. PSLA, sax-basal, sax-mid, sub4c, and supao show best performances, with a sensitivity of 100% and an AUC of more than 99%. These four views could be fully identified. A4c, a5c, a2c, and a3c were slightly misclassified among them, but the mean AUC is higher than 98%, which was also the promising results.

The confusion matrices analysis further confirmed the above results. In particular, the a2c and a3c was not easy to be classified. In general, a4c, a5c, a2c, and a3c were not misclassified with PSLA, sax-basal, sax-mid, sub4c, and supao. This result indicated the next improvement direction, i.e. the improvement on a4c, a5c, a2c and a3c, especially for a2c. The overall accuracy on independent test set was 98.15%. The proposed method could be generalized to new datasets.

The main contribution is to propose an effective method for the image classification of cardiac views. As far as we know, the obtained accuracy is the result of the state of the art. Because our data set is not large enough, we believe this problem will be improved by more data. The study confirms the potential of deep learning on ultrasound medicine, and deep learning is expected to develop a diagnostic tool for heart diseases.

## Conclusion

This paper proposed an effective deep neural network method for identifying cardiac views. The method was based on Inception V3 and introduced spatial transformer network and queue-to-excitation network, respectively. The Spatial transformer network learned the shape deformation caused by cardiac cycle and reduced intra-class differences. The squeeze-to-excitation network amplified the channel response that was effective for identification. Moreover, the structural signals among images are used based on a graph of similarity, which reduced the dependence of amount of data, and improved the generalization ability on different datasets. The work confirms the potential of deep learning on ultrasound medicine.

## Data Availability

The data that support the findings of this study are available from The First Affiliated Hospital of Xi’an Jiaotong University and Shaanxi Provincial People's Hospital but restrictions apply to the availability of these data, which were used under license for the current study, and so are not publicly available.

## Ethics approval and consent to participate

The study was approved by relevant Institutional Review Boards, and a general research authorization was obtained allowing for retrospective reviews.

## Consent for publication

Not applicable.

## Contributions

All authors read and approved the final manuscript. YM. Guo was responsible for the overall study supervision and reviewing manuscript. YH. Gao was responsible for the study design and was a major contributor in writing the manuscript. YH. Gao, Y. ZH, B. Liu were responsible for the data collection (imaging acquisition, image analysis, image interpretation, clinical and pathologic outcomes). Y.H were responsible for the statistical analysis and interpretation of the data.

## Acknowledgements

Not applicable.

